# CURE: A Pre-training Framework on Large-scale Patient Data for Treatment Effect Estimation

**DOI:** 10.1101/2022.09.09.22279776

**Authors:** Ruoqi Liu, Pin-Yu Chen, Ping Zhang

**Affiliations:** The Ohio State University; IBM Research

## Abstract

Treatment effect estimation (TEE) refers to the estimation of causal effects, and it aims to compare the difference among treatment strategies on important outcomes. Current machine learning based methods are mainly trained on labeled data with specific treatments or outcomes of interest, which can be sub-optimal if the labeled data are limited. In this paper, we propose a novel transformer-based pre-training and fine-tuning framework called **CURE** for TEE from observational data. **CURE** is pre-trained on large-scale unlabeled patient data to learn representative contextual patient representations, and then fine-tuned on labeled patient data for TEE. We design a new sequence encoding for longitudinal (or structured) patient data and we incorporate structure and time into patient embeddings. Evaluated on 4 downstream TEE tasks, **CURE** outperforms the state-of-the-art methods in terms of an average of 3.8% and 6.9% absolute improvement in Area under the ROC Curve (AUC) and Area under the Precision-Recall Curve (AUPR), and 15.7% absolute improvement in Influence function-based Precision of Estimating Heterogeneous Effects (IF-PEHE). We further demonstrate the data scalability of **CURE** and verify the results with corresponding randomized clinical trials. Our proposed method provides a new machine learning paradigm for TEE based on observational data.

## 1 Introduction

Treatment effect estimation (TEE) is to evaluate the causal effects of treatment strategies on some important outcomes, which is a crucial problem in many areas such as healthcare [12], education [9] and economics [20]. Randomized clinical trials (RCTs) are the *de-facto* gold standard for identifying causal effects through randomizing the treatment assignment and comparing the responses in different treatment groups. However, conducting RCTs is time-consuming, expensive and sometimes unethical. Observational data such as medical claims provide a promising opportunity for treatment effect estimation when RCTs are expensive or impossible to conduct.

Recently, many works have been proposed to adopt neural networks (NNs) for TEE from observational data [30, 31, 15, 7, 8, 36, 14]. Compared to classical TEE methods such as regression trees [6] or random forests [35], NN-based methods achieve better performance in handling the complex and nonlinear relationships among covariates, treatment and outcome. However, there are still some common limitations of existing TEE methods: 1) Most model designs are task-specific or data-specific so it is hard to adapt the model to a more generalized setting. 2) Existing labeled dataset often has small-scale data size, whereas training neural models requires large and high-quality labeled data for capturing inherent complex relationships of the input data.

Recently, Transformer [33] has been widely adopted as a critical and unified building block in the pre-training and fine-tuning paradigm across data modalities. The pre-trained Transformer-based models (PTMs) have become the model of choice in many deep learning domains such as natural language processing (NLP)[10, 26, 27, 3, 23] and computer vision (CV) [4, 11, 24]. The dominant approach is to pre-train on a large-scale dataset with unsupervised or self-supervised learning and then fine-tune on a smaller task-specific dataset. Nonetheless, applying this pre-training and fine-tuning paradigm to treatment effect estimation problems faces the following three major challenges: 1) encoding structured longitudinal observational patient data into sequence input; 2) lack of well-curated large-scale pre-training dataset; 3) lack of real-world downstream treatment effect estimation tasks to benchmark baselines.

In this paper, we propose a new pre-training and fine-tuning framework for **C**a**U**sal t**R**eatment **E**ffect estimation (**CURE**). As shown in Fig. 1, the large-scale structured patient data are extracted from a real-world medical claims data (MarketScan Research Databases [18]). We first encode the structured data as sequential input by chronologically flattening and aligning all observed covariates. We obtain around 3M processed unlabeled patient sequences for pre-training. And the downstream datasets with labeled treatment and outcome are created according to specific TEE tasks from established RCTs. Based on the retrospective study design and domain knowledge, we obtain 4 downstream tasks and each of them containing 10K-20K patient samples. The task is to evaluate the comparative effectiveness of two treatment effects in reducing the risk of stroke for patients with coronary artery disease (CAD). Second, we pre-train a Transformer-based model on the unlabeled data with an unsupervised learning objective to generate contextualized patient representations. To accommodate the issues of complex hierarchical structure (i.e., the patient record contains multiple visits and each visit contains multiple types of medications or diagnoses) and irregularity of the observational patient data, we propose a comprehensive embedding method to incorporate the structure and time information. Finally, we fine-tune the pre-trained model on various downstream TEE tasks.

**Figure 1:**
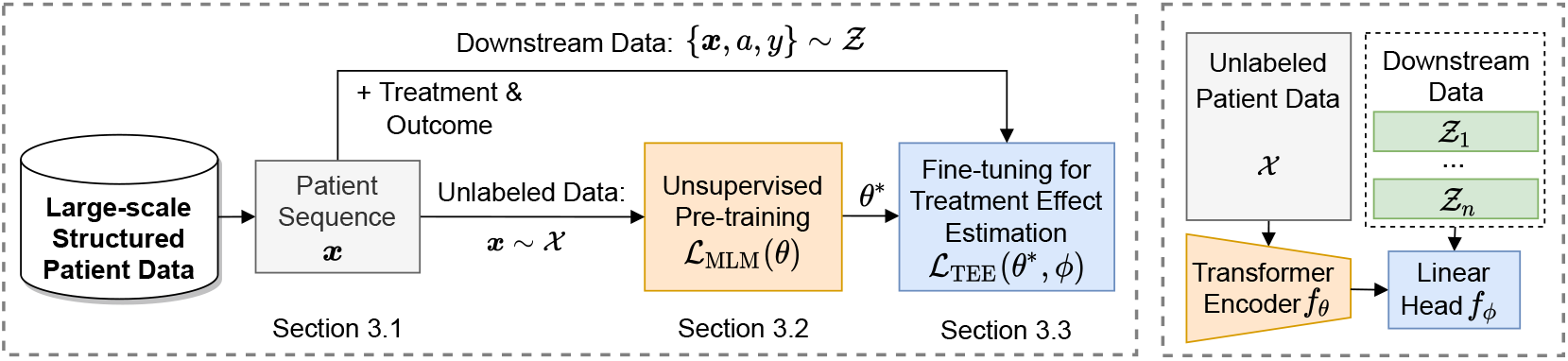
The overall pipeline of **CURE**. It mainly consists of three parts: 1) data encoding of longitudinal patient data; 2) unsupervised pre-training on unlabeled data and 3) fine-tuning on downstream labeled data for treatment effect estimation. In TEE, labels mean the studied treatment *α* and outcome *y* of the patient sequence ***x***.

We summarize our **main contributions** as follows.

- We propose **CURE**, a novel transformer-based pre-training and fine-tuning framework for TEE. We present a new patient data encoding method to encode structured observational patient data and incorporate covariate type and time into patient embeddings.
- We obtain and preprocess large-scale patient data from real-world medical claims data as our pre-training resource. We derive 4 downstream TEE tasks according to study designs and domain knowledge from established RCTs for model evaluation.
- We conduct thorough experiments and show that **CURE** yields superior performance on all downstream tasks compared to state-of-the-art TEE methods. We achieve, on average, 3.8% and 6.9% absolute improvement in AUC and AUPR respective for outcome prediction, and 15.7% absolute improvement in IF-PEHE for TEE over the best baseline among 4 tasks. We also verify the estimated treatment effects with the conclusion of corresponding RCTs.
- We further explore the effectiveness of **CURE** in several ablation studies including the proposed patient embedding, the influence of pre-training data size on downstream tasks, and the generalizability of low-resource fine-tuning data.

## 2 Background and Related Work

### Treatment effect estimation from observational data

In this paper, we are interested in observational patient data. Each patient sample consists of pre-treatment covariates *x* (i.e., historical co-medication, co-morbidities and demographics) and a treatment *a* of interest. Following the potential outcome framework [28], the potential outcome *y*_*a*_ is defined as the response to treatment *a* out of all available treatment options. Typically, we consider the comparative treatment effects of two treatments and denote two potential outcomes as *y*_1_ and *y*_0_ for simplicity.

We aim to estimate the individual-level treatment effect (ITE) as the difference between the potential outcomes under two treatment arms as *y*_1_(*x*) − *y*_0_(*x*). We are also interested in the average treatment effect (ATE) which is the average effect among the entire population, denoted as 𝔼[*y*_1_(*x*) − *y*_0_(*x*)]. In observational data, only one of the potential outcomes is available and the remaining counterfactual outcomes are missing in nature, which makes this task more difficult than classical supervised learning. To guarantee that the treatment effects are identifiable from the observational data, we follow the standard causal assumptions [21] including consistency, positivity and strong ignorability. More details of assumptions are illustrated in Appendix A.

### Deep learning for treatment effect estimation

Generally, existing NN-based methods formulate the TEE as several regression tasks (i.e., regression on potential outcomes and treatment) with different levels of information shared among the nuisance estimation tasks using representation learning. TARNet [7], for example, learns one shared representations for two potential outcomes, while SNet [7] learns five different representations on the combinations of treatment and potential outcomes. Recently, Transformer has been introduced as an encoder block for TEE [36, 14] and yields better performance compared to the state-of-the-art methods. Despite the promising results, the main limitation is that the model performance can be diminished if the labeled dataset is limited. The model trained for one particular problem or data may fail to generalize to other scenarios.

### Pre-train and fine-tune of Transformer

Since Transformer is based on a flexible architecture with few assumptions on the input data structure, it is difficult to directly train the model on small-scale data. Therefore, various pre-trained Transformer-based models (PTMs) are first pre-trained on the large-scale unlabeled data and then fine-tuned for labeled tasks at hand. PTMs learn universal and contextualized representations, which can boost various downstream tasks, and avoid developing and training a new model from scratch. Among the existing PTMs in NLP, BERT [10] is one of the most popular models. BERT [10] is pre-trained on large-scale unlabeled corpus via self-supervised pre-training tasks (i.e., masked language modeling and next sentence prediction) and fine-tuned on downstream tasks with an additional linear head. Our observational patient data are close to natural language text as they both contain sequential information. However, patient data have some unique characteristics that distinguish them from the text. Compared to the text, patient data contain a more complex hierarchical structure and time information. Therefore, existing BERT pre-training architecture can not be directly applied to modeling patient data.

## 3 CURE: A Pretraining and Fine-tuning Framework for TEE

In this section, we introduce our **CURE** framework (as shown in Fig. 1) which includes three key steps: (1) Encoding structured patient data as sequential input by aligning medications and diagnosis in each visit chronologically (Sec. 3.1), (2) Pre-training on a large-scale unlabeled patient data by minimizing the unsupervised objective and obtaining the optimized parameters *θ*\* (Sec. 3.2), and (3) Fine-tuning on a small-scale labeled downstream dataset for TEE by jointly optimizing *θ*\* and a linear head parameterized as *ϕ* (Sec. 3.3).

### 3.1 Encoding structured patient data

In this work, we focus on longitudinal observational patient data. We first introduce the data for pre-training and fine-tuning respectively. Then we illustrate how to convert structured patient data into sequential input for the Transformer encoder.

#### Pre-train data structure

The pre-training is based on large-scale unlabeled patient data. Here, to distinguish from downstream data, we denote the pre-train data as unlabeled data (*x* ∼ 𝒳), while downstream data with treatment *a* and outcome *y* as labeled data ({*x, a, y*} ∼ Ƶ). The unlabeled patient data consist of: (1) Co-medication *m*_1_, *m*_2_, …, *m*_|ℳ|_ ∈ ℳ, where |ℳ| is the number of unique medication names. (2) Co-morbidities *d*_1_, *d*_2_, …, *d*_|*𝒟*|_ ∈ 𝒟, where |𝒟| is the number of unique diagnosis codes. (3) Demographics *c*: age encoded as categorical value and gender encoded as binary value. A patient can have multiple visits {*v*_1_, …, *v*_*T*_ }, where each of visit *v*_*t*_ contains a subset of medication and diagnosis codes (*v*_*t*_ ∈ ℳ ∪ 𝒟). We denote the unlabeled patient data as 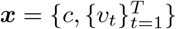. We build a medical vocabulary from all patient covariates as 𝒱 = {ℳ, 𝒟, *c*}.

#### Fine-tune data structure

The fine-tuning is based on a small-scale labeled patient data, which are not used for pre-training. Besides the co-medication, co-morbidities and demographics, the labeled patient data contain treatment *a* ∈ ℳ_task_ (i.e., can be either the target treatment or compared treatment from task-specific medication group ℳ_task_) and outcome *y*_*a*_ ∈ {0, 1} under the observed treatment *a*. In Figure. 2, we show the retrospective study design of how to construct downstream data and obtain labels for treatments and outcomes. Specially, we collect patient data from two different treatment groups for comparison. For each group of patients, all covariates are obtained from the baseline period as potential confounders, and the outcomes are obtained from the follow-up period. More illustrations of the study design can be found in Appendix C.

**Figure 2:**
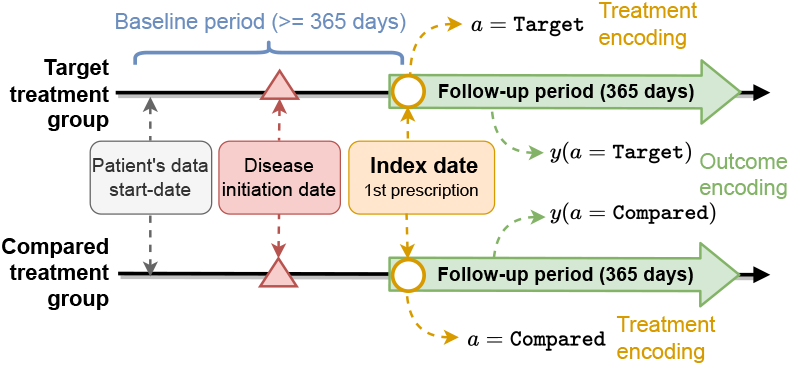
Illustration of the downstream data construction with retrospective study design. Index date refers to the first prescription of the target treatment or the compared treatment, which should be no prior to the disease initiation date. The baseline period (a.k.a, wash-out window) is no less than one year and the follow-up period as outcome observation is also one year. The treatments of interests and outcomes are obtained at the index date and follow-up period respectively.

#### Structured patient data to sequential input

As introduced above, the original patient data are recorded naturally in a hierarchical structure. Unlike natural language text, which is inherently encoded as a sequence of words, the patient data need to be preprocessed into a “sequence-like” format before sending to the Transformer encoder. As shown in Fig. 3, we flatten the structured patient data by chronologically going through each medication and diagnosis in each visit and aligning them in one sequence. Each medication or diagnosis is encoded as an individual token, which is comparable to text tokenization. The token ids are obtained from the medical vocabulary 𝒱.

**Figure 3:**
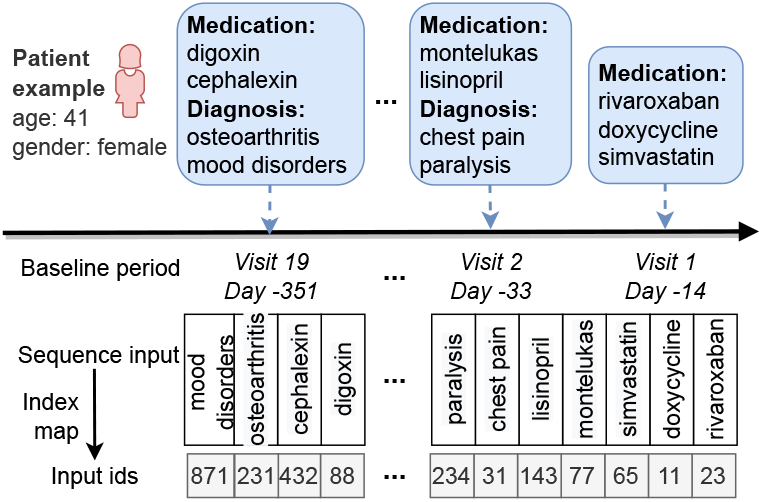
Illustration of encoding structured patient data into sequential input. The raw patient data are recorded in an hierarchical structure such that a patient contains multiple visits and each visit contains multiple medications and diagnoses. The structured data are converted into a sequence by flattening all covariates in each visit and aligning them chronologically.

### 3.2 Pre-training CURE

As shown in Fig. 4, the pre-training consists of three modules: (1) an embedding layer to convert input patient data into embedding representations, (2) Transformer encoders to generate contextualized hidden representations and (3) a final project layer for pre-training objective. More formally, given the encoded patient sequence *x* = [*x*_1_, …, *x*_*m*_, …, *x*_*T*_] as specified in Sec. 3.1, the pre-training procedure can be decomposed into the following steps:

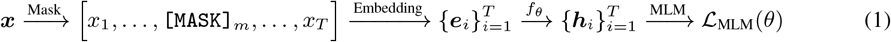

**Figure 4:**
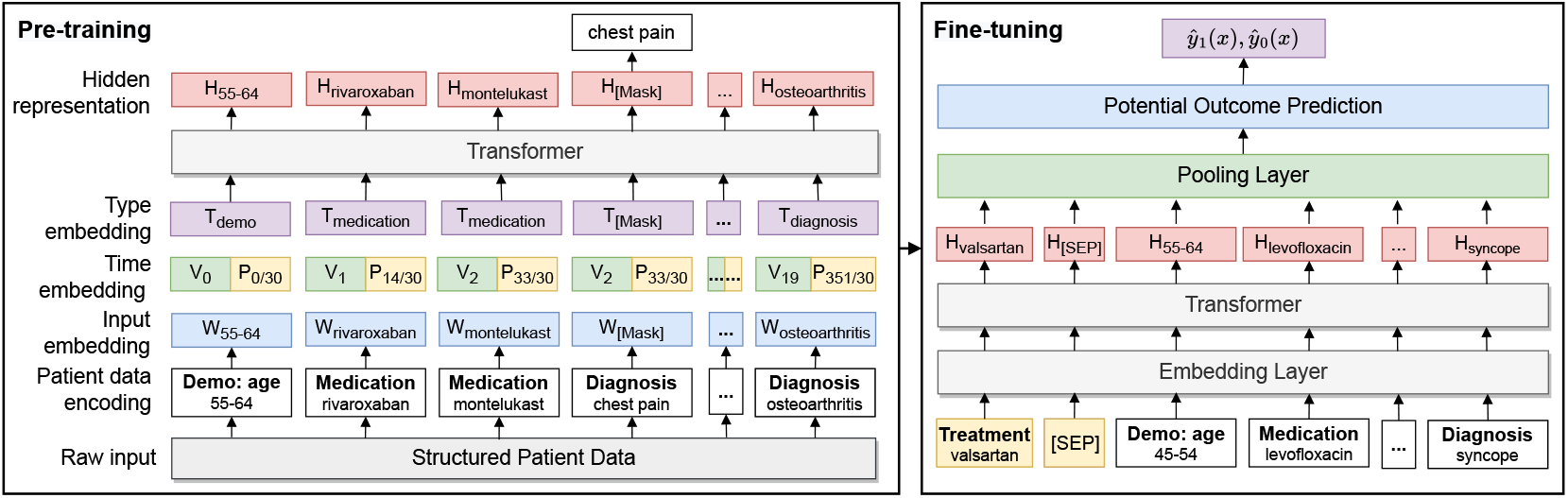
Illustration of pre-training and fine-tuning of **CURE**. The unlabeled structured patient data are first converted into a sequential input, and processed for the embedding layer and encoder. During the fine-tuning, the treatments of interest are appended for potential outcome prediction.

We randomly replace 15% of input tokens with special `[MASK]` tokens, e.g., token *x*_*m*_ in the sequence. *e*_*i*_ ∈ ℝ^*B*^ denotes the embedding representation with embedding dimension *B* generated by the comprehensive embedding layer. *h*_*i*_ ∈ ℝ^*H*^ denotes the contextualized representation with hidden dimension *H* generated by Transformer encoder *f*_*θ*_. The masked language modeling (MLM) [10] aims to predict the masked tokens *x*_*m*_ from the established vocabulary 𝒱 using hidden representation *h*_*m*_. The pre-training loss function of MLM is denoted as ℒ_MLM_(*θ*) with optimization parameters *θ*.

#### Comprehensive embedding layer

The pre-trained language models like BERT [10] have achieved great success in natural language text demonstrating strong power in modeling sequential data. Though longitudinal patient data can be deemed as a kind of sequential data when organized by the chronological order, there exist substantial and unignorable discrepancies between the text and patient data. It is hard to directly apply the existing pre-trained language model to our unique patient data. Our ablation study shows that the standard embedding design adopted in NLP (i.e., token embedding and position embedding) will be sub-optimal in our scenario (see Sec. 4.3 for more details).

Compared to the natural language text, (1) longitudinal patient data contain a more complex hierarchical structure than the text data: a patient record contains a number of visits and each visit also contains a number of different types of medical codes (i.e., medication or diagnosis). (2) The patient data are irregularly sampled (i.e., the time interval among visits is not regular) while the text data are regularly organized. As shown in Fig. 3, the visit dates are not regularly distributed along the time: the first visit happened on day 0, the second visit happened on day 14 and the third one on day 33, etc. On visit 2 (day 33), the patient received two types of codes: montelukas and lisinopril as medications, and chest pain and paralysis as diagnoses.

To accommodate the above issues of complex hierarchical structure and irregularity of the observational patient data, we propose a more comprehensive embedding layer than the original BERT [10] embedding layer by including associated code type information and time information. For each input token, the patient embedding *e*_*i*_ is obtained as:

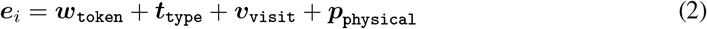

where *w*_token_ is the original input token embedding. *t*_type_ denotes the type embedding of the input token. According to our data, there are three types in total: {Demographics, Medication, Diagnosis}. The visit time embedding *v*_visit_ denotes the visit time corresponding to a visit. The physical time embedding *p*_physical_ denotes the physical time associated with the visit. Here, the physical time is measured by month (i.e., 30-day fixed window). Both visit and physical time are organized relative to the treatment index date (i.e., the absolute distance between the visit/physical time to the index date).

As an illustration, in Fig. 4, the input is a sequence of patient data containing the type and time information: rivaroxaban is from Medication type prescribed on visit 1 (day 14) and chest pain is from the Diagnosis type received on visit 2 (day 33). The input token embedding, time embedding and type embedding are integrated and used as the input to the Transformer encoder.

#### Transformer encoder and pre-training objective

We use an N-stacked Transformer as our encoding backbone as it has been a widely adopted architecture. For each single Transformer encoder block, it consists of a multi-head self-attention layer followed by a fully-connected feed-forward layer [33]. More details of the Transformer architecture are illustrated in Appendix B.

The Transformer encoder *f*_*θ*_ takes the comprehensive embedding representations as input and generates contextualized hidden representations as *f*_*θ*_(*e*). Given unlabeled patient data 𝒳, the pretraining is to minimize the MLM loss of predicting the masked token with position *m* ∈ ℳ using the input token embedding and hidden representation:

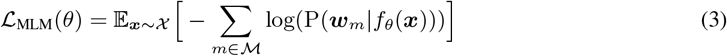

where P(*w*_*m*_|*f*_*θ*_(*x*)) is the softmax probability of the masked token over all tokens in the vocabulary.

### 3.3 Fine-tuning CURE for TEE

Given downstream labeled data {*x, a, y*} ∼ Ƶ, we fine-tune the model on different downstream TEE tasks. Here, we are interested in the comparative causal treatment effect of the target treatment over another compared treatment according to the downstream tasks. For each task, we plug in the task-specific input and outputs into **CURE**. We add a linear head *f*_*ϕ*_ to the hidden representations learned from the pre-training stage. We fully fine-tune all model parameters end-to-end by jointly updating *θ*\* obtained from optimizing Eq. 3 and a randomized *ϕ*.

Specially, we append the original input sequence with the index treatment (i.e., target treatment or compared treatment) which is separated by the special `[SEP]` token. As shown in Fig. 4, the treatment “valsartan” is appended to the original inputs to indicate that the patient is from the treatment group of “valsartan”. The model processes the new inputs through the embedding layer and the Transformer encoder with parameters initialized with *θ*\*. We use the final hidden vector corresponding to the first input token (`[CLS]`) as the pooled representation *h*_[CLS]_ from the pooling layer. We predict the potential outcomes under treatment of interest *a* via the linear head as *f*_*ϕ*_ ° *f*_*θ*_*\** (*h*_[CLS]_(*a*)). The fine-tuning objective is the binary cross entropy (BCE) of the potential outcome prediction:

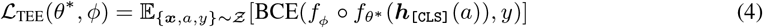

Here, only the factual outcome are used for training loss computation as the counterfactual outcomes are unavailable in the observational data. After model fine-tuning, we infer the ITE *δ* and ATE Δ as the difference between two predicted potential outcomes under the target and compared treatment:

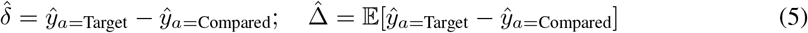

## 4 Experiments

In this section, we evaluate the proposed **CURE** from three aspects: 1) Quantitative analysis of the comparison performance with state-of-the-art TEE methods on 4 downstream tasks; 2) Qualitative analysis including the validation of the estimated treatment effects with corresponding RCTs, and self-attention feature weights visualization; 3) Ablation studies including proposed feature embedding, pre-training data size, and generalizability of low-resource fine-tuning data.

### Pre-training data

We extract patient data from MarketScan Commercial Claims and Encounters (CCAE) [18] from 2012 to 2017, which contains individual-level, de-identified healthcare claims information from employers, health plans and hospitals. In this paper, we evolve patients who have ever been diagnosed with coronary artery disease (CAD) as our disease cohort. The definition of CAD is in Appendix C. After conducting data preprocessing and study design, we obtain 2,955,399 patient sequences for pre-training. We obtain 9,435 medical codes including 282 diagnosis codes (i.e., we map the original ICD-9/10 billing codes into Clinical Classifications Software [CCS] [16]) and 9,153 medication codes (i.e., we map medications based on generic names from RED BOOK [19]).

### Downstream tasks

As the ground truth treatment effects are not available in observational data, we use RCTs as the gold standard to verify our results. We focus on CAD-related RCTs which study the comparative effectiveness of two treatments for reducing the risk of stroke after CAD. We first collect all available Phase 2 and Phase 3 RCTs with CAD as disease name and stroke as disease outcome from https://clinicaltrials.gov/. Stroke is selected because it is commonly used as the primary outcome measurement in various CAD studies and it is well-defined in observational data. Then we select completed RCTs with published results. Finally, we end up with 4 RCTs that meet all the above criteria. We derive corresponding downstream tasks from our data based on the study design as specified in Fig. 2. More details of screening RCTs are in Appendix C.

### Baselines

We compare **CURE** with 8 neural network models for TEE, including state-of-the-art methods. For models designed for continuous outcomes with mean square error (MSE) as a training objective, we change the objective function to binary cross entropy for consistency. All the baselines are only trained on downstream data and are summarized below:

- *TARNet* [30] is the first model proposed for TEE via neural networks. The model predicts the potential outcomes based on balanced representations among treated and controlled groups.
- *DragonNet* [31] jointly optimizes treatment prediction and potential outcome prediction. The model first learns shared representations given the input data and then does prediction tasks via a three-head neural network: one for treatment prediction and two for potential outcomes.
- *DR-CFR* [15] learns disentangled representations for counterfactual regression. The assumption is that the observed covariates can be disentangled into three components: only contributing to treatment selection, only contributing to outcome predication, and both.
- *TNet* [7] is a neural network based T-learner (Two-learner) [22]. T-learner is a kind of meta-learners that decomposes the TEE into two sub-regression problems. TNet fits two neural models as base learners to estimate the outcome under treatment and control.
- *SNet* [7] is based on DR-CFR [15] and assumes that the observed covariates can be disentangled into five components by considering two potential outcomes separately.
- *FlexTENet* [8] incorporates the idea of inductive bias for shared structure of two potential outcomes into TEE. The model adaptively learns what to share between the potential outcome functions.
- *TransTEE* [36] is a recently proposed Transformer-based TEE model. The covariates and treatments are encoded via a Transformer and aggregated for outcome prediction via a cross-attention layer.
- *Base Model* directly trains on the downstream datasets using the same architecture as **CURE**.

### Metrics

We evaluate the factual prediction performance using the standard classification metrics: Area under the ROC Curve (AUC) and Area under the Precision-Recall Curve (AUPR). We evaluate the counterfactual prediction performance using the influence function-based precision of estimating heterogeneous effects (IF-PEHE) [1], which helps to benchmark TEE methods when the ground truth effects are not available. Compared to the widely adopted precision of estimating heterogeneous effects (PEHE) that measures the mean squared error between estimated treatment effects and true treatment effects, IF-PEHE measures the mean squared error between estimated treatment effects and approximated true treatment effects. The output of the IF-PEHE metric is a numeric value and the lower the better. More details of this metric are in Appendix C.

### Implementation details

Our pre-training uses the BERT_*base*_ architecture [10] with 768 hidden size, 12 attention heads, 12 layer Transformer and 3072 intermediate size. The maximum input sequence length is 256. The pre-training is conducted on 3 NVIDIA GeForce RTX 2080 Ti 11GB GPUs with a batch size of 96. We train our model using the adaptive moment estimation (Adam) optimizer, with an initial learning rate of 1*e* − 4 and learning rate warmup in the first 10% training steps. During the fine-tuning, the learning rate is 5*e* − 5 without learning rate warmup. We fine-tune the model on each task for 2 epochs. The downstream data are randomly split into training, validation and test sets with percentages of 90%, 5%, 5% respectively. All results are reported on the test sets. More implementation details are mentioned in Appendix C. The code of our proposed **CURE** is available in Supplementary material.

### 4.1 Quantitative analysis

#### Comparison with state-of-the-art methods

Table 1 shows the performance of factual outcome prediction (measured by AUC and AUPR) and TEE (measured by IF-PEHE) on four different downstream tasks. We compare **CURE** with the state-of-the-art TEE methods and report the results under 20 random runs. We observe that the proposed **CURE** has more than 3.8%, 6.9% and 15.7% respective average AUC, AUPR and IF-PEHE improvement over the best baseline on these tasks. The results illustrate the promise and effectiveness of our proposed pre-training and fine-tuning methodology for TEE. Among all baselines, SNet [7] and TransTEE [36] generally perform better than others. Notably, even without pre-training, the base model of **CURE** attains similar performance as the best baseline, which suggests the effectiveness of our architecture and data encoding designs.

**Table 1:**
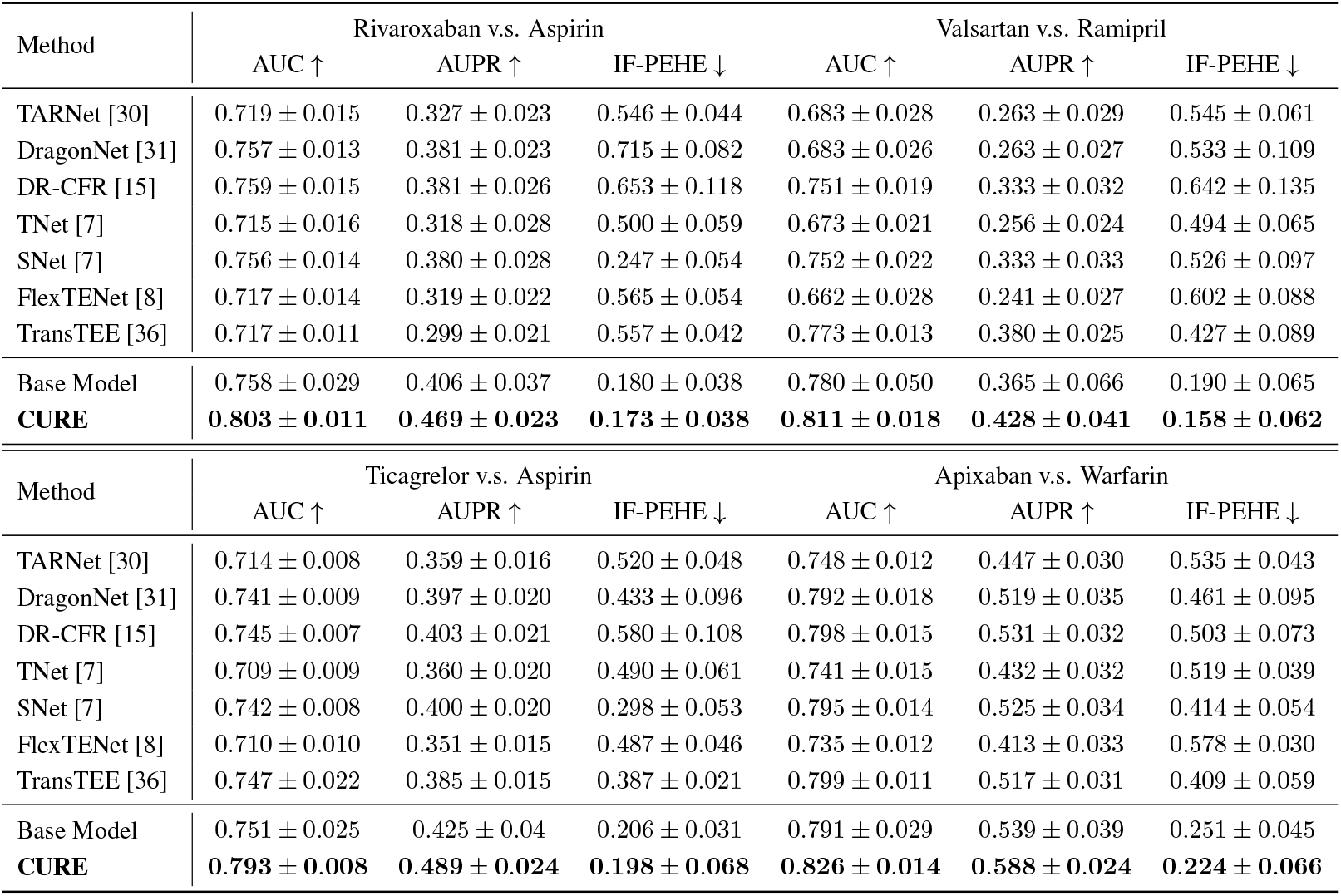
Comparison with state-of-the-art methods on four downstream datasets. The results are the average and standard deviation over 20 runs.

| Method | Rivaroxaban v.s. Aspirin |  |  | Valsartan v.s. Ramipril |  |  |
| --- | --- | --- | --- | --- | --- | --- |
| | AUC ↑ | AUPR ↑ | IF-PEHE ↓ | AUC ↑ | AUPR ↑ | IF-PEHE ↓ |
| TARNet [30] | 0.719 ± 0.015 | 0.327 ± 0.023 | 0.546 ± 0.044 | 0.683 ± 0.028 | 0.263 ± 0.029 | 0.545 ± 0.061 |
| DragonNet [31] | 0.757 ± 0.013 | 0.381 ± 0.023 | 0.715 ± 0.082 | 0.683 ± 0.026 | 0.263 ± 0.027 | 0.533 ± 0.109 |
| DR-CFR [15] | 0.759 ± 0.015 | 0.381 ± 0.026 | 0.653 ± 0.118 | 0.751 ± 0.019 | 0.333 ± 0.032 | 0.642 ± 0.135 |
| TNet [7] | 0.715 ± 0.016 | 0.318 ± 0.028 | 0.500 ± 0.059 | 0.673 ± 0.021 | 0.256 ± 0.024 | 0.494 ± 0.065 |
| SNet [7] | 0.756 ± 0.014 | 0.380 ± 0.028 | 0.247 ± 0.054 | 0.752 ± 0.022 | 0.333 ± 0.033 | 0.526 ± 0.097 |
| FlexTENet [8] | 0.717 ± 0.014 | 0.319 ± 0.022 | 0.565 ± 0.054 | 0.662 ± 0.028 | 0.241 ± 0.027 | 0.602 ± 0.088 |
| TransTEE [36] | 0.717 ± 0.011 | 0.299 ± 0.021 | 0.557 ± 0.042 | 0.773 ± 0.013 | 0.380 ± 0.025 | 0.427 ± 0.089 |
| Base Model | 0.758 ± 0.029 | 0.406 ± 0.037 | 0.180 ± 0.038 | 0.780 ± 0.050 | 0.365 ± 0.066 | 0.190 ± 0.065 |
| **CURE** | **0.803 ± 0.011** | **0.469 ± 0.023** | **0.173 ± 0.038** | **0.811 ± 0.018** | **0.428 ± 0.041** | **0.158 ± 0.062** |

| Method | Ticagrelor v.s. Aspirin |  |  | Apixaban v.s. Warfarin |  |  |
| --- | --- | --- | --- | --- | --- | --- |
| | AUC ↑ | AUPR ↑ | IF-PEHE ↓ | AUC ↑ | AUPR ↑ | IF-PEHE ↓ |
| TARNet [30] | 0.714 ± 0.008 | 0.359 ± 0.016 | 0.520 ± 0.048 | 0.748 ± 0.012 | 0.447 ± 0.030 | 0.535 ± 0.043 |
| DragonNet [31] | 0.741 ± 0.009 | 0.397 ± 0.020 | 0.433 ± 0.096 | 0.792 ± 0.018 | 0.519 ± 0.035 | 0.461 ± 0.095 |
| DR-CFR [15] | 0.745 ± 0.007 | 0.403 ± 0.021 | 0.580 ± 0.108 | 0.798 ± 0.015 | 0.531 ± 0.032 | 0.503 ± 0.073 |
| TNet [7] | 0.709 ± 0.009 | 0.360 ± 0.020 | 0.490 ± 0.061 | 0.741 ± 0.015 | 0.432 ± 0.032 | 0.519 ± 0.039 |
| SNet [7] | 0.742 ± 0.008 | 0.400 ± 0.020 | 0.298 ± 0.053 | 0.795 ± 0.014 | 0.525 ± 0.034 | 0.414 ± 0.054 |
| FlexTENet [8] | 0.710 ± 0.010 | 0.351 ± 0.015 | 0.487 ± 0.046 | 0.735 ± 0.012 | 0.413 ± 0.033 | 0.578 ± 0.030 |
| TransTEE [36] | 0.747 ± 0.022 | 0.385 ± 0.015 | 0.387 ± 0.021 | 0.799 ± 0.011 | 0.517 ± 0.031 | 0.409 ± 0.059 |
| Base Model | 0.751 ± 0.025 | 0.425 ± 0.04 | 0.206 ± 0.031 | 0.791 ± 0.029 | 0.539 ± 0.039 | 0.251 ± 0.045 |
| **CURE** | **0.793 ± 0.008** | **0.489 ± 0.024** | **0.198 ± 0.068** | **0.826 ± 0.014** | **0.588 ± 0.024** | **0.224 ± 0.066** |

### 4.2 Qualitative analysis

#### Validate with RCT conclusion

As the ground truth treatment effects are not available in observational data, we further evaluate the estimated treatment effects with corresponding ground truth RCTs. In Table 4.2, we show the confidence intervals of estimated effects under 20 runs and RCT conclusions of each downstream task.

**Table 2:** Comparison of the estimated treatment effects with corresponding ground truth RCT. The estimated effects are shown in 95% confidence intervals (CI) under 20 bootstrap runs. The RCT conclusions are obtained from published articles.

| Target v.s. Compared | Estimated Effect (CI) | P value | Generated Hypothesis | RCT Conclusion |
| --- | --- | --- | --- | --- |
| Rivaroxaban v.s. Aspirin | [-0.009, 0.006] | 0.452 | No significant difference | No significant difference [2] |
| Valsartan v.s. Ramipril | [-0.003, 0.014] | 0.103 | No significant difference | No significant difference [25] |
| Ticagrelor v.s. Aspirin | [0.022, 0.040] | 6e-14 | T. is less effective than A. | No significant difference [29] |
| Apixaban v.s. Warfarin | [-0.039, -0.002] | 4e-4 | A. is more effective than W. | A. is more effective than W. [13] |

We use the direct difference to estimate the treatment effects [17]. The results can be interpreted as two potential conclusions: 1) The target treatment is significantly more effective than the compared treatment in reducing the risk of the outcome if the upper bound of the confidence interval is lower than zero. (2) The target is not significantly more effective than the compared treatment if the confidence interval covers zero (i.e., no significant difference) or the lower bound is higher than zero (i.e., the compared treatment is more effective than the target treatment). As we can see, our estimated treatment effects are mostly consistent with each corresponding RCT conclusion. Though the generated hypothesis and RCT conclusion are not exactly the same for the third pair (Ticagrelor v.s. Aspirin), they both indicate that there is no significant reduced treatment effect of the target treatment over the compared treatment. The results demonstrate that our proposed **CURE** successfully identifies correct treatment effects using only observational patient data.

#### Self-attention visualization

The self-attention mechanism of the Transformer enables the exploration of interaction among input covariates and provides a potential interpretation of the prediction results. We use a Transformer visualization tool called bertviz [34] to help visualize learned attention weights. We show the visualization results of some patient samples in Appendix D.

### 4.3 Ablation studies

#### Effect of embedding layer

We evaluate the effect of proposed time embedding (visit time and physical time) and type embedding respectively. As shown in Fig. 5, the model with both time and type embedding generally performs better than the other two embedding ablations. Especially, incorporating time embedding yields larger performance improvement than the type embedding. This indicates that the proposed embedding method is better than the standard embedding method and time information plays a more important role in TEE than the type information.

**Figure 5:**
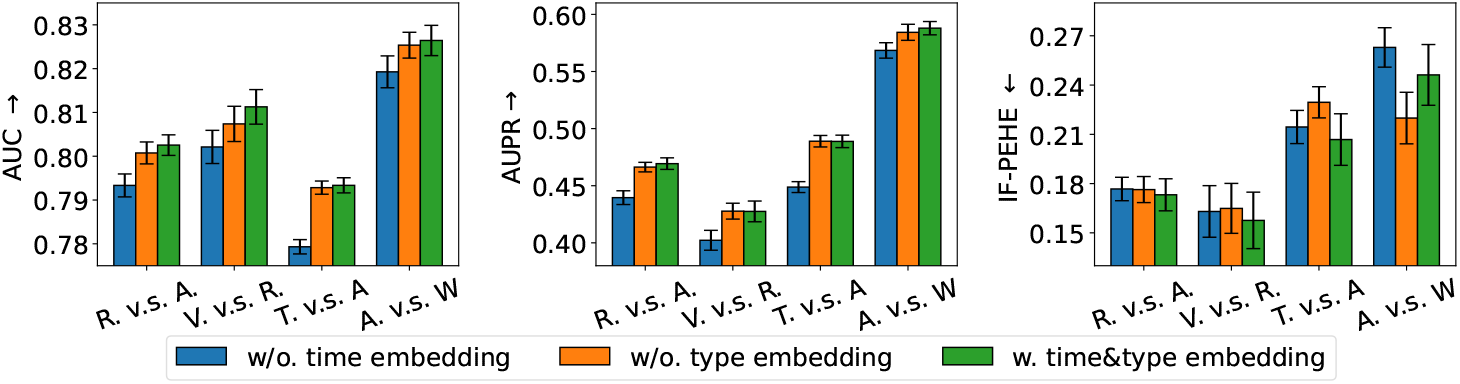
The effect of different embedding layer designs on four downstream tasks.

#### Effect of downstream data size

We demonstrate the model’s effectiveness on the low resource of downstream data in Fig. 6. The plots show the model performance with different fractions of labeled downstream data. Generally, given only 5% 10% labeled data, the **CURE** achieves comparable performance to the Base Model which is trained on the fully labeled data. Specifically, the performance gains are large when given a small fraction of labeled data (1%-5%) and the curve tends to gently increase after the fraction is larger than 10%. With increased data size, the performance gradually achieves the upper bound of fine-tuning on fully labeled data. The results demonstrate that unsupervised pre-training benefits low-resource downstream tasks even when only a limited number of labeled data are available for fine-tuning. More results of other metrics are in Appendix D.

**Figure 6:**
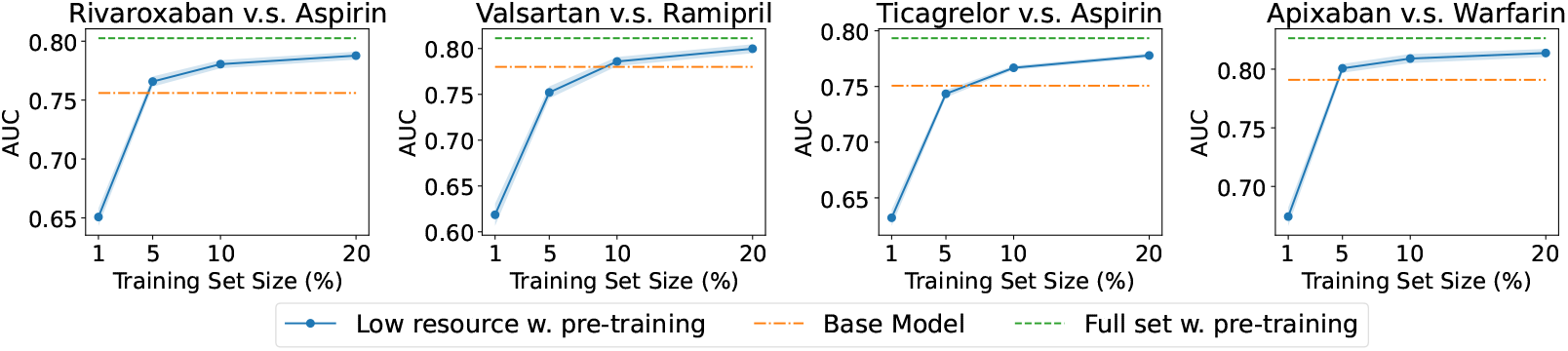
The effect of low resource in fine-tuning datasets on four downstream tasks with different fractions of labeled training set (x-axes). The results are the average of 20 runs.

#### Effect of pre-training data size

We further explore the effect of pre-training data volume on the performance of downstream tasks. In Fig. 7, we show the AUC given different fractions of pretraining data. Here, 0% training set size denotes the Base Model, which is trained on the downstream data from scratch. Generally, the performance improves with the increase of pre-train data. The results indicate that pre-training is beneficial for downstream tasks by learning contextualized patient representations from large-scale unlabeled patient data. Other metrics are show in Appendix D.

**Figure 7:**
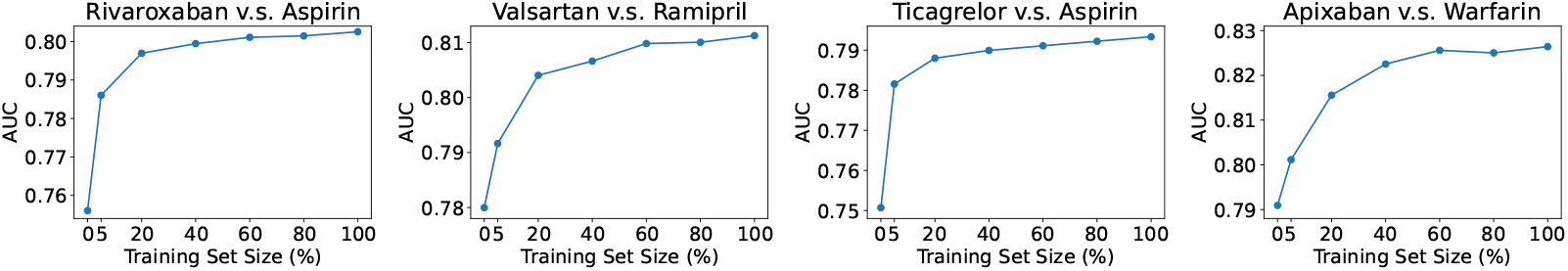
The effect of pre-training data volume on four downstream tasks (average of 20 runs).

## 5 Conclusion

In this paper, we study the problem of TEE from observational data. We propose a new transformer-based TEE framework called **CURE**, which adopts the pre-training and fine-tuning paradigm. **CURE** is pre-trained on a large-scale unlabeled patient data and then fine-tuned on labeled patient data for TEE. We convert the structured patient data into sequence and design a new sequence encoding method to encode the structure and time into a comprehensive patient embedding. Thorough experiments show that pre-training significantly boosts the TEE performance on 4 downstream tasks compared to state-of-the-art methods. We further demonstrate the data scalability of **CURE** and verify the results with corresponding published RCTs. One promising application of our model is to help generate useful hypotheses of treatment effects and serve as a complementary tool to standard RCTs, e.g., exploring the new uses of existing drugs. Future works could be done to improve the model performance by engaging more patient data from diverse disease cohorts as pre-training data.

### Ethical consideration

The observational data used in the paper are from IBM MarketScan Research Database, which is fully HIPAA-compliant de-identified, have very minimal risk of the potential for loss of privacy. Moreover, Per the DUAs with MarketScan, all users to access the data will need to take full research, ethics, and compliance training courses and be covered by IRBs. Thus, potential privacy and security risk would be eliminated and/or mitigated.

## Data Availability

The experimental data can be obtained from IBM MarketScan Research Databases.

https://www.ibm.com/products/marketscan-research-databases

## References

[1] Ahmed Alaa and Mihaela Van Der Schaar. Validating causal inference models via influence functions. In International Conference on Machine Learning, pages 191–201. PMLR, 2019.

[2] Sonia S Anand, Jackie Bosch, John W Eikelboom, Stuart J Connolly, Rafael Diaz, Peter Widimsky, Victor Aboyans, Marco Alings, Ajay K Kakkar, Katalin Keltai, et al. Rivaroxaban with or without aspirin in patients with stable peripheral or carotid artery disease: an international, randomised, double-blind, placebo-controlled trial. The Lancet, 391(10117):219–229, 2018.

[3] Tom Brown, Benjamin Mann, Nick Ryder, Melanie Subbiah, Jared D Kaplan, Prafulla Dhariwal, Arvind Neelakantan, Pranav Shyam, Girish Sastry, Amanda Askell, et al. Language models are few-shot learners. Advances in neural information processing systems, 33:1877–1901, 2020.

[4] Nicolas Carion, Francisco Massa, Gabriel Synnaeve, Nicolas Usunier, Alexander Kirillov, and Sergey Zagoruyko. End-to-end object detection with transformers. In European conference on computer vision, pages 213–229. Springer, 2020.

[5] Tianqi Chen and Carlos Guestrin. Xgboost: A scalable tree boosting system. In Proceedings of the 22nd acm sigkdd international conference on knowledge discovery and data mining, pages 785–794, 2016.

[6] Hugh A Chipman, Edward I George, and Robert E McCulloch. Bart: Bayesian additive regression trees. The Annals of Applied Statistics, 4(1):266–298, 2010.

[7] Alicia Curth and Mihaela van der Schaar. Nonparametric estimation of heterogeneous treatment effects: From theory to learning algorithms. In International Conference on Artificial Intelligence and Statistics, pages 1810–1818. PMLR, 2021.

[8] Alicia Curth and Mihaela van der Schaar. On inductive biases for heterogeneous treatment effect estimation. Advances in Neural Information Processing Systems, 34, 2021.

[9] Rajeev H Dehejia and Sadek Wahba. Causal effects in nonexperimental studies: Reevaluating the evaluation of training programs. Journal of the American statistical Association, 94(448):1053–1062, 1999.

[10] Jacob Devlin, Ming-Wei Chang, Kenton Lee, and Kristina Toutanova. Bert: Pre-training of deep bidirectional transformers for language understanding. arXiv preprint 1810.04805, 2018.

[11] Alexey Dosovitskiy, Lucas Beyer, Alexander Kolesnikov, Dirk Weissenborn, Xiaohua Zhai, Thomas Unterthiner, Mostafa Dehghani, Matthias Minderer, Georg Heigold, Sylvain Gelly, et al. An image is worth 16×16 words: Transformers for image recognition at scale. arXiv preprint 2010.11929, 2020.

[12] Thomas A Glass, Steven N Goodman, Miguel A Hernán, and Jonathan M Samet. Causal inference in public health. Annual review of public health, 34:61–75, 2013.

[13] Christopher B Granger, John H Alexander, John JV McMurray, Renato D Lopes, Elaine M Hylek, Michael Hanna, Hussein R Al-Khalidi, Jack Ansell, Dan Atar, Alvaro Avezum, et al. Apixaban versus warfarin in patients with atrial fibrillation. New England Journal of Medicine, 365(11):981–992, 2011.

[14] Zhenyu Guo, Shuai Zheng, Zhizhe Liu, Kun Yan, and Zhenfeng Zhu. Cetransformer: Casual effect estimation via transformer based representation learning. In Chinese Conference on Pattern Recognition and Computer Vision (PRCV), pages 524–535. Springer, 2021.

[15] Negar Hassanpour and Russell Greiner. Learning disentangled representations for counterfactual regression. In International Conference on Learning Representations, 2019.

[16] Healthcare Cost and Utilization Project. Clinical Classifications Software (CCS). Available at www.hcup-us.ahrq.gov/toolssoftware/ccs/ccs.jsp.

[17] Miguel Angel Hernán. A definition of causal effect for epidemiological research. Journal of Epidemiology & Community Health, 58(4):265–271, 2004.

[18] IBM MarketScan Research Databases. Available at https://www.ibm.com/products/marketscan-research-databases.

[19] IBM Micromedex RED BOOK. Available at https://www.ibm.com/products/micromedex-red-book.

[20] Guido W Imbens. Nonparametric estimation of average treatment effects under exogeneity: A review. Review of Economics and statistics, 86(1):4–29, 2004.

[21] Guido W Imbens and Donald B Rubin. Causal inference in statistics, social, and biomedical sciences. Cambridge University Press, 2015.

[22] Sören R Künzel, Jasjeet S Sekhon, Peter J Bickel, and Bin Yu. Metalearners for estimating heterogeneous treatment effects using machine learning. Proceedings of the national academy of sciences, 116(10):4156–4165, 2019.

[23] Yinhan Liu, Myle Ott, Naman Goyal, Jingfei Du, Mandar Joshi, Danqi Chen, Omer Levy, Mike Lewis, Luke Zettlemoyer, and Veselin Stoyanov. Roberta: A robustly optimized bert pretraining approach. arXiv preprint 1907.11692, 2019.

[24] Niki Parmar, Ashish Vaswani, Jakob Uszkoreit, Lukasz Kaiser, Noam Shazeer, Alexander Ku, and Dustin Tran. Image transformer. In International Conference on Machine Learning, pages 4055–4064. PMLR, 2018.

[25] Marc A Pfeffer, Brian Claggett, Eldrin F Lewis, Christopher B Granger, Lars Køber, Aldo P Maggioni, Douglas L Mann, John JV McMurray, Jean-Lucien Rouleau, Scott D Solomon, et al. Angiotensin receptor– neprilysin inhibition in acute myocardial infarction. New England Journal of Medicine, 385(20):1845–1855, 2021.

[26] Alec Radford, Karthik Narasimhan, Tim Salimans, and Ilya Sutskever. Improving language understanding by generative pre-training. 2018.

[27] Alec Radford, Jeffrey Wu, Rewon Child, David Luan, Dario Amodei, Ilya Sutskever, et al. Language models are unsupervised multitask learners. OpenAI blog, 1(8):9, 2019.

[28] Donald B Rubin. Causal inference using potential outcomes: Design, modeling, decisions. Journal of the American Statistical Association, 100(469):322–331, 2005.

[29] Sigrid E Sandner, Heribert Schunkert, Adnan Kastrati, Dominik Wiedemann, Martin Misfeld, Andreas Boening, Ulrich Tebbe, Bernd Nowak, Jan Stritzke, Guenther Laufer, et al. Ticagrelor monotherapy versus aspirin in patients undergoing multiple arterial or single arterial coronary artery bypass grafting: insights from the ticab trial. European Journal of Cardio-Thoracic Surgery, 57(4):732–739, 2020.

[30] Uri Shalit, Fredrik D Johansson, and David Sontag. Estimating individual treatment effect: generalization bounds and algorithms. In International Conference on Machine Learning, pages 3076–3085. PMLR, 2017.

[31] Claudia Shi, David Blei, and Victor Veitch. Adapting neural networks for the estimation of treatment effects. Advances in neural information processing systems, 32, 2019.

[32] John W Stanifer, Sean D Pokorney, Glenn M Chertow, Stefan H Hohnloser, Daniel M Wojdyla, Samira Garonzik, Wonkyung Byon, Ziad Hijazi, Renato D Lopes, John H Alexander, et al. Apixaban versus warfarin in patients with atrial fibrillation and advanced chronic kidney disease. Circulation, 141(17):1384–1392, 2020.

[33] Ashish Vaswani, Noam Shazeer, Niki Parmar, Jakob Uszkoreit, Llion Jones, Aidan N Gomez, Łukasz Kaiser, and Illia Polosukhin. Attention is all you need. Advances in neural information processing systems, 30, 2017.

[34] Jesse Vig. A multiscale visualization of attention in the transformer model. In Proceedings of the 57th Annual Meeting of the Association for Computational Linguistics: System Demonstrations, pages 37–42, Florence, Italy, July 2019. Association for Computational Linguistics.

[35] Stefan Wager and Susan Athey. Estimation and inference of heterogeneous treatment effects using random forests. Journal of the American Statistical Association, 113(523):1228–1242, 2018.

[36] Yi-Fan Zhang, Hanlin Zhang, Zachary C Lipton, Li Erran Li, and Eric P Xing. Can transformers be strong treatment effect estimators? arXiv preprint 2202.01336, 2022.

