## Supplementary Material for "CURE: A Pre-training Framework on Large-scale Patient Data for Treatment Effect Estimation"

### Appendix

#### A Causal Assumptions

We follow the standard causal assumptions [5] to help guarantee that the treatment effects are identifiable from the observational data.

**Assumption 1 (Consistency)** *The potential outcome under the treatment $a$ equals to the observed outcome if the actual treatments is $a$.*

**Assumption 2 (Positivity)** *Given the observational data of the history, if the the probability $P(a=1|\boldsymbol{x}) \neq 0$, then the probability of receiving treatment $0$ or $1$ is positive, i.e., $0 < P(A=a|X=\boldsymbol{x}) < 1$, for all $a \in \mathcal{A}$ and $\boldsymbol{x} \in \mathcal{X}$.*

**Assumption 3 (Strong Ignorability)** *Given the observational data of the history, the treatment assignment is independent of the potential outcome, i.e., $Y(A=a) \perp\!\!\!\perp A|X=\boldsymbol{x}$, for all $a \in \mathcal{A}$.*

Assumption 1 is fundamental to the potential outcome framework used to define counterfactuals and infer treatment effects. Essentially, this assumption requires that the treatment specified in the study must be precise enough that any variation within the treatment specification will not lead to a different outcome. Assumption 2 implies that all patients may receive the treatment whatever their observed covariates. Otherwise, it is impossible to derive the counterfactuals for patients who do not have any chance of being in the other treatment group. Assumption 3 states that the potential outcomes are independent of treatment assignment given the set of observed covariates. This assumption guarantees that the treatment effects are identifiable given the treatment, outcome and observed covariates as: $\mathbb{E}[Y(A=1) - Y(A=0)] = \mathbb{E}_{\boldsymbol{x} \in \mathcal{X}}[\mathbb{E}[Y|A=1,\boldsymbol{x}] - \mathbb{E}[Y|A=0,\boldsymbol{x}]]$.

#### B Transformer Architecture

For each single Transformer encoder block, it consists of a multi-head self-attention layer followed by a fully-connected feed-forward layer [6]. The multi-head attention is the most crucial part which can be calculated as,

$$\begin{aligned} \text{MultiHead}(\boldsymbol{h}) &= \text{Concat}(\text{head}_1, \ldots, \text{head}_h)W^O; \\ \text{head}_i &= \text{Attention}(\boldsymbol{h}W_i^Q, \boldsymbol{h}W_i^K, \boldsymbol{h}W_i^V) \\ \text{Attention}(Q,K,V) &= \text{Softmax}(\frac{QK^T}{\sqrt{d}})V \end{aligned} \tag{1}$$

where $\boldsymbol{h} \in \mathbb{R}^{d \times d_{\text{model}}}$ denotes the hidden representations and $d$ is the input sequence length. $W_i^Q \in \mathbb{R}^{d_{\text{model}} \times d}$, $W_i^K \in \mathbb{R}^{d_{\text{model}} \times d}$, $W_i^V \in \mathbb{R}^{d_{\text{model}} \times d}$, $W^O \in \mathbb{R}^{nd \times d_{\text{intermediate}}}$ are learnable parameter matrices. $d = d_{\text{model}}/n$ and $n$ is the number of attention heads. We show the detailed model configuration in Fig. A1.

```
"attention_probs_dropout_prob": 0.1,
"classifier_dropout": null,
"hidden_act": "gelu",
"hidden_dropout_prob": 0.1,
"hidden_size": 768,
"initializer_range": 0.02,
"intermediate_size": 3072,
"layer_norm_eps": 1e-12,
"max_physical_time_embeddings": 13,
"max_position_embeddings": 512,
"max_visit_time_embeddings": 361,
"model_type": "bert",
"num_attention_heads": 12,
"num_hidden_layers": 12,
"pad_token_id": 0,
"position_embedding_type": "absolute",
"time_embedding": true,
"torch_dtype": "float32",
"transformers_version": "4.17.0",
"type_vocab_size": 5,
"use_cache": true,
"vocab_size": 9452
```

Figure A1: Model configuration.

#### C Additional Details on Experimental Setups

**Pre-training data.** The pre-training data are obtained from MarketScan Commercial Database [4], which consists of medical and drug data from employers and health plans for over 215 million individuals. In this study, we focus on CAD as the studied disease and stroke as the outcome. The definitions of CAD and stroke are shown in Table A1 and Table A2 respectively.

Table A1: The definition of coronary artery disease (CAD) from observational health data.

| | |
|---|---|
| Reference (PMID) | 16159046, 26524702, 28008010 |
| Criteria | A history of coronary revascularization in the EHR<br>Or, history of acute coronary syndrome, ischemic heart disease, or exertional angina |
| Diagnostic codes | ICD-9 codes:<br>410* to 414*<br>ICD-10 codes:<br>The best approximation are the following codes:<br>I20* Angina pectoris<br>I21* Acute myocardial infarction<br>I22* Subsequent ST elevation (STEMI) and non-ST elevation (NSTEMI) myocardial infarction<br>I23* Certain current complications following ST elevation (STEMI) and non-ST elevation (NSTEMI) myocardial infarction (within the 28 day period)<br>I24* Other acute ischemic heart diseases<br>I25* Chronic ischemic heart disease |

Table A2: The definition of stroke from observational health data

| | |
|---|---|
| Reference (PMID) | 29202795 |
| Diagnostic codes | ICD-9 codes:<br>V12.54,<br>438.0–438.9<br>ICD 10 codes:<br>Z86.73<br>I60-I69<br>subarachnoid hemorrhage (I60);<br>intracerebral hemorrhage (I61);<br>cerebral infarction (I63);<br>and other transient cerebral ischemic attacks and related syndromes and transient cerebral ischemic attack (unspecified) (G458 and G459) |

**Downstream tasks.** We demonstrate the flowchart for RCT extraction in Fig. A2. All RCTs are extracted from `https://clinicaltrials.gov/`. We start from 1,593 CAD-related RCTs with stroke as the outcome and end up with 4 RCTs that satisfy all the above criteria. We have included all those 4 RCTs for downstream task construction.

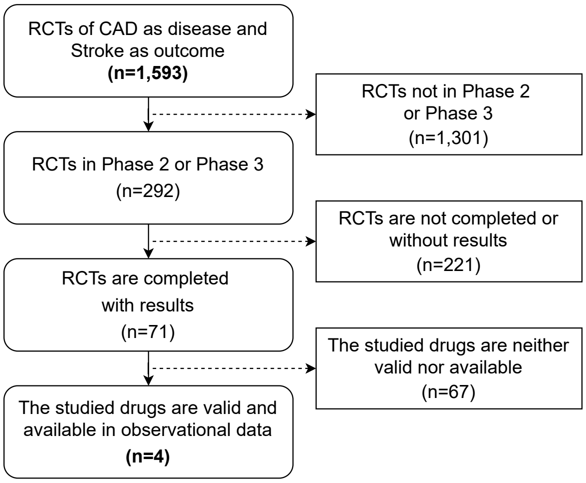


Figure A2: The data flow for RCT extraction. The downstream tasks constructed based on the extracted RCTs.

Table A3: The statistics of the downstream dataset.

| Target v.s. Compared | Rivaroxaban v.s. Aspirin | Valsartan v.s. Ramipril | Ticagrelor v.s. Aspirin | Apixaban v.s. Warfarin |
|---|---|---|---|---|
| # of patients (Target; Compared) | 26340 (9569; 16771) | 12850 (7306; 5544) | 29248 (12477; 16771) | 18187 (6701; 11486) |
| Female (%) | 30.4 | 32.4 | 27.1 | 31.8 |
| Age (group) on index date | 55-64 | 55-64 | 55-64 | 55-64 |
| Patients with stroke (%) | 13.7 | 11.9 | 18.9 | 16.7 |
| Average # of visits per patient | 83.4 | 74.0 | 70.7 | 97.1 |
| Average # of codes per patient | 182.3 | 157.2 | 152.0 | 215.9 |

**Pre-training and fine-tuning data preparation.** The pre-training is based on large-scale unlabeled patient data, and the fine-tuning is based on small-scale labeled patient data, which are not used for pre-training. We first construct 4 datasets for downstream tasks according to the study design of related randomized clinical trial (RCT). The patients who satisfy the eligibility criteria of the RCT are included in each dataset respectively (see more details of the study design in Fig. 2). Then we construct the unlabeled pre-training data based on all the remaining patients who are not included in any of the 4 downstream datasets. Therefore, the same patients do not appear simultaneously in both the pre-training stage and the fine-tuning stage.

**Evaluation metrics.** As the true treatment effects are not available in real-world data, we use the influence function-based precision of estimating heterogeneous effects (IF-PEHE) [1] for model evaluation. Following the same experimental setup, we calculate IF-PEHE as,

- Step 1: Train two XGBoost [2] classifiers for potential outcome prediction denoted by $\mu_0$ and $\mu_1$, where $\mu_a = P(y_a = 1|X = x)$ using the training set $\mathcal{Z}_{\text{train}}$. Then calculate the plug-in estimation $\widetilde{T} = \mu_1 - \mu_0$ Train a XGBoost [2] classifier propensity score function (i.e., the probability of receiving treatment) $\widetilde{\pi} = P(a = 1|X = x)$.
- Step 2: Given the estimated treatment effect $\hat{T}(x_i)$ on the test set $\mathcal{Z}_{\text{test}}$, calculate the IF-PEHE with the influence function $\hat{l}$ as,

$$\begin{aligned} \text{IF-PEHE} &= \sum_{x_i \in \mathcal{Z}_{\text{test}}} [(\hat{T}(x_i) - \widetilde{T}(x_i))^2 + \hat{l}(x_i)] \\ \hat{l}(x) &= (1 - B)\widetilde{T}^2(x) + By(\widetilde{T}(x) - \hat{T}(x)) - W(\widetilde{T}(x) - \hat{T}(x))^2 + \hat{T}^2(x) \end{aligned} \tag{2}$$

where $W = (a - \widetilde{\pi}(x))$, $B = 2a(a - \widetilde{\pi}(x))C^{-1}$, $C = \widetilde{\pi}(x)(1 - \widetilde{\pi}(x))$.

**Implementation details.** The pre-training model architecture follows the $\text{BERT}_{\text{base}}$ [3] and most hyper-parameters remain the same as default setting. The detailed hyperparameters setup is shown in Table A4 for pre-training , and Table A5 for fine-tuning. With 3 NVIDIA GeForce RTX 2080 Ti 11GB GPUs, the pre-training takes about 20 hours with current setup. We have provided all code in supplemental material.

Table A4: Hyperparameters used in pre-training.

| Parameters | **CURE** |
|---|---|
| Maximum Steps | 100K |
| Initial Learning Rate | 1e-4 |
| Batch Size | 96 |
| Warm-Up Steps | 10K |
| Sequence Length | 256 |
| Dropout | 0.1 |

Table A5: Hyperparameters search space and optimal parameters used for fine-tuning.

| Parameters | Search Space | Optimal Value |
|---|---|---|
| Maximum Epochs | {1,2,3,4,5} | 2 |
| Initial Learning Rate | {1e-5, 3e-5, 5e-5} | 5e-5 |
| Batch Size | {16, 32, 64} | 32 |
| Sequence Length | 256 | 256 |
| Fixed Window Length | 30 | 30 |
| Baseline Window | {90, 180, 360, 720} | 360 |
| Dropout | 0.1 | 0.1 |

Table A6: The parameter size of the proposed method and baselines.

| Method | Model parameters |
|---|---|
| TARNet [9] | 2M |
| DragonNet [8] | 2M |
| DR-CFR [12] | 3M |
| TNet [10] | 4M |
| SNet [10] | 3M |
| FlexTENet [11] | 3M |
| TransTEE [13] | 7M |
| CURE | 93M |

Table A7: The influence of weight ($\alpha$) associated with the discriminator in DragonNet to the model performance on the Valsartan v.s. Ramipril dataset (random seed =42).

| $\alpha$ | AUC | AUPR | IF-PEHE |
|---|---|---|---|
| 0.2 | 0.677 | 0.304 | 0.768 |
| 0.4 | 0.679 | 0.308 | 0.689 |
| 0.6 | 0.680 | 0.310 | 0.660 |
| 0.8 | 0.682 | 0.312 | 0.644 |
| 1.0 | 0.683 | 0.314 | 0.643 |
| 1.2 | 0.679 | 0.315 | 0.593 |
| 1.4 | 0.681 | 0.317 | 0.584 |
| 1.6 | 0.682 | 0.318 | 0.589 |
| 1.8 | 0.683 | 0.319 | 0.586 |
| 2.0 | 0.685 | 0.321 | 0.595 |
| CURE | 0.805 | 0.428 | 0.161 |

### D Additional Experimental Results

**Visualization.** The self-attention mechanism of the Transformer enables the exploration of interaction among input covariates. As an example, we show the attention weights of a patient from Apixaban treatment group of Apixaban v.s. Warfarin study in Fig. A3. Different colors denote the attention heads and there are 12 heads in total. The medications and diagnosis codes highlighted in the figure are the most related features to the outcome prediction and treatment effect estimation. For example, Amiodarone is an antiarrhythmic medication used to treat and prevent a number of types of cardiac dysrhythmias including atrial fibrillation [1]. A study [7] shows that apixaban is superior to warfarin in preventing stroke in patients with atrial fibrillation. Those attention weights could be used to analyze the treatment effects in some subgroups that characterized by the attended feature set.

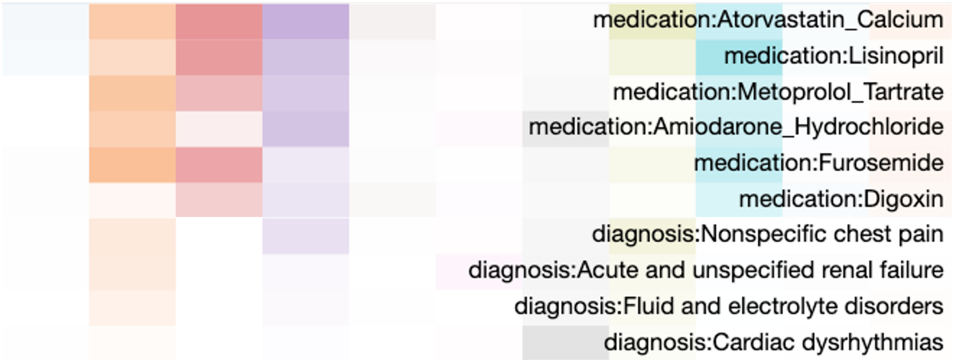


Figure A3: The visualization of Top 10 attention weights associated with the special token `[CLS]` of a patient from Apixaban treatment group.

**Ablation studies.** We show the results of two ablation studies (Sec. 4 **Ablation studies.**) in terms of AUPR scores in Fig. A4 and Fig. A5, respectively. For the effect of low resource in fine-tuning, our model can achieve comparable performance as measured by AUPR to the Base Model with only around 5%~10% labeled downstream data. For the effect of pre-trainign data size, the model gradually yields better performance when the pre-train data size increases.

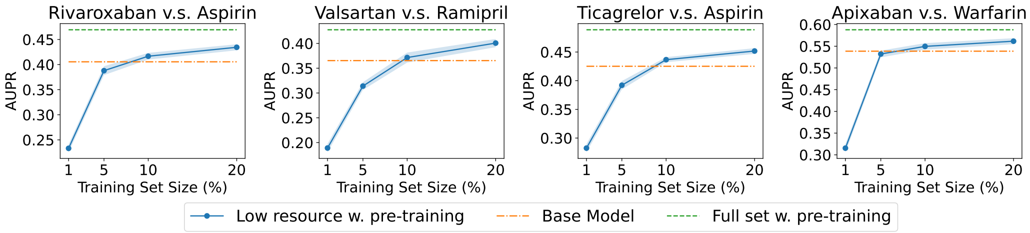


Figure A4: The effect of low resource in fine-tuning datasets on four downstream tasks with different fractions of labeled training set (x-axes). The results are the average of 20 runs.

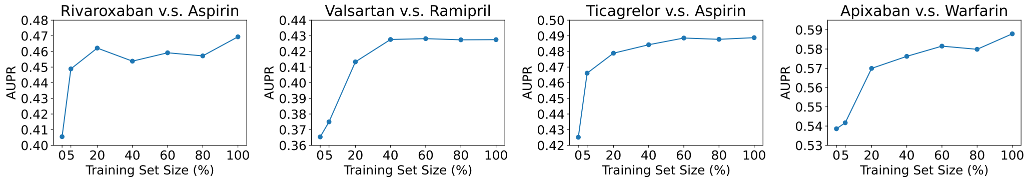


Figure A5: The effect of pre-training data volume on four downstream tasks (average of 20 runs).

[1] https://www.drugs.com/monograph/amiodarone.html

**Evaluation on non-random assignment.** We show the t-SNE visualization of learned patient representations of treatment and control group respectively (see Fig. A6). The visualization of our model (CURE) demonstrates that the distribution variance between two groups is marginal and the non-random assignment issue is alleviated.

We further adapt the design in DragonNet [8] in our model's fine-tuning stage. Specifically, we add an additional prediction head for propensity score estimation and modify the loss function to incorporate both outcome prediction and propensity prediction. We compare the new model (CURE+propensity) with the proposed CURE model on 4 downstream tasks respectively. As shown in Table A8, the performance of these two models is comparable in terms of both factual prediction and treatment effect estimation.

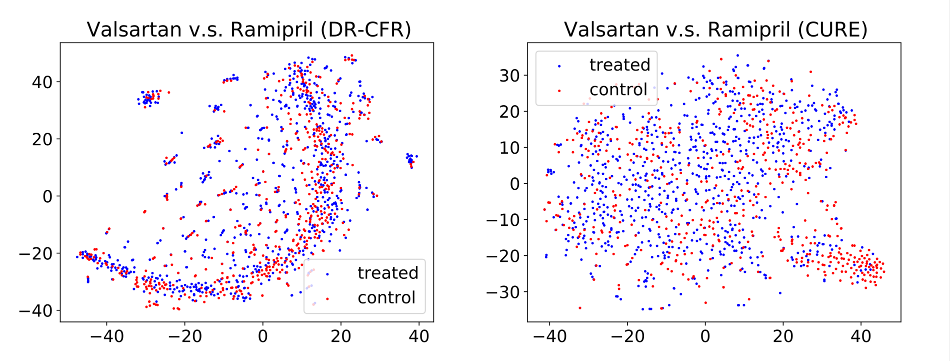


Figure A6: The t-SNE visualization of learned patient representations on Valsartan v.s. Ramipril dataset.

Table A8: Comparison of the proposed model (CURE) and a new model based on CURE but adapting propensity module in DragonNet [8] (CURE + propensity) on 4 downstream tasks (random seed=42).

| | Rivaroxaban v.s. Aspirin | | | Valsartan v.s. Ramipril | | |
|---|---|---|---|---|---|---|
| Method | AUC | AUPR | IF-PEHE | AUC | AUPR | IF-PEHE |
| CURE | 0.786 | 0.419 | 0.186 | 0.805 | 0.428 | 0.161 |
| CURE+propensity | 0.789 | 0.427 | 0.178 | 0.804 | 0.428 | 0.156 |

| | Ticagrelor v.s. Aspirin | | | Apixaban v.s. Warfarin | | |
|---|---|---|---|---|---|---|
| Method | AUC | AUPR | IF-PEHE | AUC | AUPR | IF-PEHE |
| CURE | 0.807 | 0.502 | 0.211 | 0.838 | 0.597 | 0.182 |
| CURE+propensity | 0.806 | 0.505 | 0.239 | 0.844 | 0.602 | 0.253 |

**Additional evaluation with RCT conclusion.** We evaluate the treatment effects estimated by all the baselines and conduct the same hypothesis testing. As shown in Table A9 below, our method correctly generates 3 (out of 4) RCT conclusions that match the ground truth RCT conclusions while the best baselines only identify 2 (out of 4) RCT conclusions.

Table A9: Comparison of the estimated treatment effects with corresponding ground truth RCT of all methods.

| Method | Rivaroxaban v.s. Aspirin Estimated Effect (CI) | P value | Match RCT Conclusion? | Valsartan v.s. Ramipril Estimated Effect (CI) | P value | Match RCT Conclusion? |
|---|---|---|---|---|---|---|
| TARNet [9] | [0.066, 0.095] | 5.678e-10 | No | [-0.037, -0.003] | 0.026 | No |
| DragonNet [8] | [0.18, 0.236] | 5.979e-12 | No | [0.03, 0.07] | 4.681e-05 | No |
| DR-CFR [12] | [0.13, 0.183] | 2.783e-10 | No | [0.002, 0.04] | 0.033 | No |
| TNet [10] | [0.041, 0.07] | 2.509e-07 | No | [-0.038, -0.001] | 0.039 | No |
| SNet [10] | [-0.002, 0.008] | 0.231 | Yes | [-0.051, -0.026] | 3.168e-06 | No |
| FlexTENet [11] | [0.064, 0.108] | 1.529e-07 | No | [-0.079, -0.035] | 3.184e-05 | No |
| TransTEE [13] | [-0.013, -0.002] | 0.018 | No | [-0.019, 0.034] | 0.420 | Yes |
| CURE | [-0.009, 0.006] | 0.452 | Yes | [-0.003, 0.014] | 0.103 | Yes |

| Method | Ticagrelor v.s. Aspirin Estimated Effect (CI) | P value | Match RCT Conclusion? | Apixaban v.s. Warfarin Estimated Effect (CI) | P value | Match RCT Conclusion? |
|---|---|---|---|---|---|---|
| TARNet [9] | [0.064, 0.101] | 2.861e-08 | No | [-0.006, 0.028] | 0.207 | No |
| DragonNet [8] | [-0.013, 0.01] | 0.821 | Yes | [0.018, 0.056] | 6.284e-04 | No |
| DR-CFR [12] | [-0.068, -0.029] | 4.915e-05 | No | [-0.026, -0.002] | 0.047 | Yes |
| TNet [10] | [0.046, 0.069] | 6.474e-09 | No | [0.009, 0.023] | 2.329e-04 | No |
| SNet [10] | [0.005, 0.016] | 4.398e-04 | No | [-0.046, -0.017] | 2.112e-04 | Yes |
| FlexTENet [11] | [0.045, 0.068] | 5.243e-09 | No | [0.012, 0.042] | 0.001 | No |
| TransTEE [13] | [-0.014, -0.009] | 0.0216 | No | [-0.027, -0.002] | 0.027 | Yes |
| CURE | [0.022, 0.040] | 5.982e-14 | No | [-0.039, -0.002] | 4e-04 | Yes |

**Semi-synthetic experiment.** We generate a semi-synthetic dataset based on real patient data obtained from the MarketScan data. Specifically, we simulate treatment assignment $a$ and potential outcome $y$ using pre-treatment covariates $\boldsymbol{x}$ (i.e., historical co-medication, co-morbidities and demographics). The treatment assignment is simulated by $a|\overline{x} \sim \text{Bernoulli}(\text{Sigmoid}(s^T\overline{x} + m))$, where $s \sim \mathcal{N}(0^{|\mathcal{V}|}, 0.1 \cdot I)$, $|\mathcal{V}|$ is the cardinality of medical feature vocabulary, $m \sim \mathcal{N}(0, 0.1)$, $\overline{x}$ denotes the aggregation of all historical covariates. The outcome is simulated by $y|\overline{x}, a \sim \text{Bernoulli}(\text{Sigmoid}(w^T\overline{x} + \beta a + n))$, where $w \sim \mathcal{N}(0^{|\mathcal{V}|}, 0.1 \cdot I)$, $\beta \sim \mathcal{N}(0, 1), n \sim \mathcal{N}(0, 0.1)$.

As we have all potential outcomes under both treatment and control arms available in the semi-synthetic data, the model performance is evaluated with Precision of Estimating Heterogeneous Effects (PEHE), which measures the root mean square error between the true treatment effect and estimated treatment effect. The comparison results from the semi-synthetic dataset are shown in Table A10. The proposed model CURE yields the best performance among all the baselines. We will add the results for the semi-synthetic dataset in revision.

Table A10: Comparison with state-of-the-art methods on semi-synthetic Valsartan v.s. Ramipril dataset. The results are the average and standard deviation over 20 runs.

| Method | PEHE |
|---|---|
| TARNet [9] | $0.768 \pm 0.012$ |
| DragonNet [8] | $0.759 \pm 0.015$ |
| DR-CFR [12] | $0.714 \pm 0.014$ |
| TNet [10] | $0.784 \pm 0.017$ |
| SNet [10] | $0.776 \pm 0.022$ |
| FlexTENet [11] | $0.791 \pm 0.014$ |
| TransTEE [13] | $0.689 \pm 0.012$ |
| CURE | $\mathbf{0.596 \pm 0.010}$ |

#### References


[1] Ahmed Alaa and Mihaela Van Der Schaar. Validating causal inference models via influence functions. In *International Conference on Machine Learning*, pages 191–201. PMLR, 2019.

[2] Tianqi Chen and Carlos Guestrin. Xgboost: A scalable tree boosting system. In *Proceedings of the 22nd acm sigkdd international conference on knowledge discovery and data mining*, pages 785–794, 2016.

[3] Jacob Devlin, Ming-Wei Chang, Kenton Lee, and Kristina Toutanova. Bert: Pre-training of deep bidirectional transformers for language understanding. *arXiv preprint arXiv:1810.04805*, 2018.

[4] IBM MarketScan Research Databases.
Available at `https://www.ibm.com/products/marketscan-research-databases`.

[5] Guido W Imbens and Donald B Rubin. *Causal inference in statistics, social, and biomedical sciences*. Cambridge University Press, 2015.

[6] Ashish Vaswani, Noam Shazeer, Niki Parmar, Jakob Uszkoreit, Llion Jones, Aidan N Gomez, Łukasz Kaiser, and Illia Polosukhin. Attention is all you need. *Advances in neural information processing systems*, 30, 2017.

[7] John W Stanifer, Sean D Pokorney, Glenn M Chertow, Stefan H Hohnloser, Daniel M Wojdyla, Samira Garonzik, Wonkyung Byon, Ziad Hijazi, Renato D Lopes, John H Alexander, et al. Apixaban versus warfarin in patients with atrial fibrillation and advanced chronic kidney disease. *Circulation*, 141(17):1384–1392, 2020.

[8] Claudia Shi, David Blei, and Victor Veitch. Adapting neural networks for the estimation of treatment effects. *Advances in neural information processing systems*, 32, 2019.

[9] Uri Shalit, Fredrik D Johansson, and David Sontag. Estimating individual treatment effect: generalization bounds and algorithms. In *International Conference on Machine Learning*, pages 3076–3085. PMLR, 2017.

[10] Alicia Curth and Mihaela van der Schaar. Nonparametric estimation of heterogeneous treatment effects: From theory to learning algorithms. In *International Conference on Artificial Intelligence and Statistics*, pages 1810–1818. PMLR, 2021.

[11] Alicia Curth and Mihaela van der Schaar. On inductive biases for heterogeneous treatment effect estimation. *Advances in Neural Information Processing Systems*, 34, 2021.

[12] Negar Hassanpour and Russell Greiner. Learning disentangled representations for counterfactual regression. In *International Conference on Learning Representations*, 2019.

[13] Yi-Fan Zhang, Hanlin Zhang, Zachary C Lipton, Li Erran Li, and Eric P Xing. Can transformers be strong treatment effect estimators? *arXiv preprint arXiv:2202.01336*, 2022.
